# A Longitudinal Study of Real-World Physical Activity Assessed Using Cut—point Free Metrics, Mobility Capacity, and Mobility Perception Among Older Adults Recovering from Proximal Femoral Fracture

**DOI:** 10.64898/2026.08.07.26359957

**Authors:** Hananeh Younesian, David Singleton, Beatrix Vereijken, Judith Garcia-Aymerich, Lynn Rochester, Martin A. Berge, Monika Engdal, Joren Buekers, Sarah Koch, Jorunn L. Helbostad, Paula Alvarez, Carl-Philipp Jansen, Jochen Klenk, Kamiar Aminian, Anisoara Paraschiv-Ionescu, Clemens Becker, Brian Caulfield

## Abstract

Physical activity (PA) is measured objectively through daily wearable monitoring and mobility capacity tests, and subjectively via patient reported outcomes (mobility perception). This study investigated longitudinal changes in, and relationships between, different measures of PA among older adults recovering from proximal femoral fracture (PFF). Participants (N=201) were classified into four groups by time since surgery at baseline (T1) and followed over two assessments (T2, T3). They wore an accelerometer for 7 consecutive days. Daily PA was measured using cut-point free metrics including Average Acceleration, Intensity Gradient, and intensity of the most active accumulated X minutes (MX: M1-M90). Mobility capacity and perception of participants were evaluated using clinical tests (e.g., 6-minute Walking Test (6MinWT)) and questionnaires (Late-Life Function and Disability Instrument (LLFDI)). MX metrics, particularly M1-M15, increased significantly across the first three groups with higher sensitivity in group 1 (p<0.001).

Distance covered during the 6MinWT increased significantly (p<0.01). Three of the seven LLFDI’s domains showed the largest significant changes. Overall, sustained, moderate-strong positive correlations were observed between the clinical tests, LLFDI, and short-duration MX metrics in group 3 and 4 at T1, and across all participants at T2 and T3. Thus, MX metrics (M1-M15) can reveal change for daily PA intensities, especially among PFF groups in early recovery groups at T1 and reached the late stage at follow-ups. Clinicians may focus on specific LLFDI’s domains to maximize assessment efficiency. The direct links between mobility capacity, perceived mobility, and short-duration MX metrics indicate the potential of these metrics to monitor patients remotely.

## I. INTRODUCTION

THE global incidence of Proximal Femoral Fractures (PFF), is forecasted to reach almost 20 million annually by 2050 [1], [2]. A PFF is a major injury which results in deteriorated daily physical activity (PA) instantly. Moreover, it is associated with several deleterious health outcomes and high mortality levels [3], [4], [5]. Early mobilization after surgery has been identified as one of the modifiable factors that can impact mortality rate [6]. Therefore, longitudinal mobility assessment comes with a high clinical importance.

Nowadays, clinicians increasingly consider using wearable devices like accelerometers to assess daily PA, evaluate clinical and surgical outcomes, and monitor changes alongside their traditional assessments to better understand patients’ daily PA level and their needs [7]. Wearable devices are able to provide objective, long-term data gathered in an individual’s habitual environment [8], [9]. In contrast, traditional mobility capacity (e.g., 6-minute Walking Test (6MinWT), 4-meter Walking Test (4MWT), Short Physical Performance Battery (SPPB)) or mobility perception tests (patient reported outcomes (PROs): e.g., Late-Life Function and Disability Instrument (LLFDI)) assess within controlled environments (i.e., laboratory, hospital) [10], [11], [12], [13]. Thus, these are susceptible to the Hawthorne effect, ceiling or floor effects, and recall bias [14], [15].

The traditional cut-point based method for analyzing daily PA intensity using wearable device is limited by protocol- and population-dependent (making data comparison difficult), arbitrary boundaries, and inability to classify PA for individuals who don’t reach established thresholds [16], [17]. Consequently, researchers are shifting towards recent advancements in accelerometer data processing, cut-point free metrics, driven by raw acceleration which avoid predefined cut-points entirely [18],[19]. Cut-point free metrics facilitate the measurement of individuals’ PA profiles to deliver valuable insights about their daily volume and intensity of PA. Cut-point free metrics include Average Acceleration (AvAcc) which represents volume or overall PA, Intensity Gradient (IG) which describes the overall intensity distribution of PA, and MX metrics, the minimum acceleration of the most active accumulated X-minutes through the day [20], [21], [22], [23]. Larger AvAcc and MX metrics represent a more intense overall daily PA in specified durations, and are expressed in milligravitational units (mg) [22], [24]. The IG represents the negative curvilinear relationship between PA intensity and time accumulated at that intensity during 24 hours [21], [25], [22]. A more negative IG (steeper drop) reflects a lower amount of time accumulated at mid-range and higher intensities, while a less negative IG (shallower drop) reflects more time spread across the intensity range [19], [21], [25].

Higher AvAcc and IG have been associated with longer life expectancy in a cohort of middle-aged adults [20]. Another study on post-menopausal women and adults with type-2 diabetes showed a positive association between higher IG, bone health and physical function [26]. Thus, it seems that the association between cut-point free metrics, clinical tests, and PROs can provide an overview of daily PA intensity and volume for clinicians. However, age, demographic characteristics, disease stage, and recovery phase can impact these associations. For example, a recent cross-sectional study among PFF patients revealed a strong and positive relationship between short duration of MX metrics (M1-M30) and mobility capacity outcomes, particularly in patients at a later stage of recovery [27].

To date and to our knowledge, no study measured cut-point free metrics longitudinally among a PFF cohort to evaluate daily PA and assess its association with mobility capacity and perception tests. Accordingly, the objectives of this study were twofold: 1) to measure the changes in daily PA (AvAcc, IG, MX metrics), mobility capacity (6MinWT, 4MWT, SPPB), mobility perception (LLFDI), and quantify their sensitivity to changes; 2) to assess the longitudinal association between the cut-point free metrics and common clinical tests among PFF patients at different stages.

## II. Methods

### A. Design

This is a longitudinal study using baseline (T1) assessment, 6-month (T2), and 12-month (T3) follow-ups of the PFF cohort as part of the Mobilise-D Clinical Validation Study (version: V.7.1, Clinical Trial Registry Number: ISRCTN12051706).

### B. Participants

Of the 395 participants with PFF who attended and completed their sensor assessment at T1, 201 participants underwent both T2 and T3 follow-up evaluations. Information on ethical approval, recruitment procedures, and inclusion and exclusion criteria of the Mobilise-D CVS are published elsewhere [28], [29]. Table A (appendix) provides a comparison of age, demographic characteristics, and relevant clinical outcomes between included at T1 and subsequently excluded due to lack of follow-ups.

To allow comparison among participants across three assessments, participants were divided into four groups based on the number of days between the surgery date and T1 assessment: group 1) acute group (≤14 days post-surgery), group 2) post-acute group (15 to 42 days post-surgery), group 3) extended recovery group (43 to 182 days post-surgery), and group 4) long-term recovery group (≥183 post-surgery) [29]. At T2 follow-up, the recovery stage of group 1 turned to either extended recovery or long-term recovery groups. The recovery stage of all participants reached log-term recovery at T3 follow-up. Each group was evaluated separately across the three assessments.

### C. Tasks and Procedures

#### 1 Daily PA Assessment

Daily PA was monitored using a single wearable device (AX6, Axivity Ltd., Newcastle upon Tyne, UK) directly attached to the lower back of participants via a custom-designed adhesive patch. Participants were requested to wear the device continuously for 24 hours per day for 7 consecutive days. For this study, we used the triaxial accelerometer data of the device (sampling frequency 100 Hz, a range of ±8 g, and a resolution of 1 mg). The device has a battery life of up to 14 days.

#### 2 Mobility Capacity Assessment

Mobility capacity was measured using the 6MinWT, 4MWT, and SPPB test in the clinic [28], [29]. These tests were carried out in a straight hallway and on a flat surface. The 6MinWT measures the distance an individual can walk over a 6-minute period [30]. The participants were asked to walk back and forth around two cones that were 20 m apart, as fast as they could. During the 6MinWT, participants were allowed walking aids and rest, if needed [30]. Of note, participants in the acute group at T1 could not perform the 6MinWT. For the 4MWT, participants walked 4 meters at both of their comfortable and their self-selected speed. It was repeated twice and the fastest trial was recorded as the maximum self-selected walking speed. The SPPB has three parts: a three-stage balance test, five times sit-to-stand test, and the 4MWT test. Each part is scored from 0 to 4 points; the total score ranges from 0 (worst) to 12 (best) [30].

#### 3 Mobility Perception Assessment

Mobility perception was evaluated using the LLFDI questionnaire during approximately 30-minute clinical interviews. The LLFDI consists of two main components (Function and Disability) divided into seven domains [12]. The Upper, Basic Lower, and Advanced Lower Extremities domains of the Function component are used to measure the difficulty to perform discrete actions or activities [12], [13]. The Instrumental Role and Management Role domains of the Disability component are related to the Limitations Dimension, assessing the limitations in performing activities at home/in the community and limitations in organization or management of social tasks that involve minimal mobility, respectively. The Personal and Social Role domains of the Disability component are related to the Frequency Dimension, assessing the frequency of various personal tasks and the frequency of performing various social/community tasks, respectively. To simplify clinical interpretation, raw LLFDI scores were scaled ranging from 0 to 100, with higher scores representing better function with less limitations [12]. Of note, LLFDI scores were not available for the acute group at T1.

### D. Accelerometer Processing

To analyze the raw (.csv) accelerometer data, we used the open source GGIR package (version 3.1.4) of the statistical programming language R (version 4.4.1). Data processing steps included autocalibration, identification of non-wear periods, and calculation of the average dynamic acceleration corrected for gravity (Euclidean Norm minus 1 g, ENMO) over 5-second epochs and reported in mg units (1 mg = 0.00981 m/s2). Non-wear time detection has been explained previously [27]. Participants were excluded if their accelerometer data totalized less than 3 valid days (defined as >14 h per day) [22], [23]. The GGIR configuration has been described elsewhere [27].

Finally, the following cut-point free metrics were extracted for each valid day and then averaged over the available days:

AvAcc (mg): arithmetic average of dynamic acceleration accumulated in a 24-hour day. It is a combination of PA time and intensity and reflects overall volume of PA (GGIR argument: do.enmo = TRUE) [21], [23].

IG: negative curvilinear relationship between intensity and time accumulated at that intensity during the 24-hour day, reflecting intensity distribution. The IG is always negative, representing drop-in time accumulated as intensity increases. A steeper drop (more negative IG) means little time accumulated at midrange and higher physical intensities, whereas shallower drop (less negative IG) displays more time spread across the intensity range (GGIR argument: iglevels = TRUE) [22].

Eight different MX metrics (M1, M2, M5, M10, M15, M30, M60, M90 (mg)): minimum acceleration value above which the most active X-minutes are accumulated. The active minutes do not need to be in bouts and can be accumulated in any way across the day (GGIR argument: i.e., for M5 qlevels = c(1440-5)/1440) [25].

Without weighting, AvAcc, IG, and MX metrics were averaged across valid days.

### E. Statistical Analysis

#### 1 Primary Analysis

The primary goal was to measure the changes in daily PA (AvAcc, IG, MX metrics), mobility capacity (6MinWT, 4MWT, SPPB), mobility perception (LLFDI), and to quantify their sensitivity to changes. Therefore, repeated-measures ANOVA or Friedman tests were used to evaluate changes across three assessments (T1, T2, T3) among cut-point free metrics (AvAcc, IG, M1-M90), 6MinWT, 4MWT, SPPB, and LLFDI’s domains for each recovery group separately. Paired t-tests, or Wilcoxon rank tests if normality was not met, were applied to measure changes of 6MinWT and LLFDI domains for group 1 at T2 and T3. Changes in participants’ characteristics between T1 and T3 were analyzed using the same approach

Accounting for the exploratory nature of these analyses, Benjamini & Hochberg’s method was applied to adjust p-values, by controlling the false discovery rate at 0.05 [31], [32]. Effect size thresholds for each statistical test were categorized negligible, small, medium, and large according to Cohen 1992 to evaluate sensitivity to change [33].

#### 2 Secondary Analysis

The secondary outcome was to assess the longitudinal relationship between cut-point free metrics, mobility capacity, and mobility perception tests among PFF individuals at different stages of recovery. Spearman rank correlation was used to determine the association between the cut-point free metrics and the clinical tests for each assessment and each recovery group separately. The correlation strength (r_s_) was categorized as follows: negligible (r_s_: < 0.1), small (r_s_: 0.1<0.3), medium (r_s_: 0.3 to < 0.5) and large (r_s_: ≥ 0.5) [34]. To quantify sensitivity to change and precision of the observed associations, effect size statistics and confidence intervals (CI) were reported for the measurement of change and correlation tests, respectively [35].

All statistical analyses were conducted using SPSS (version 29.0.1; Armonk, NY, USA) and R (version 4.4.1).

## III. RESULTS

The participants’ characteristics are presented in Table I. The total size of the sample analyzed was 201; the median age of groups 1 to 4 was 76, 80, 77, and 78 years, respectively. The BMI of the participants was similar across the four recovery groups and did not change significantly from T1 to T3.

**TABLE I.** Participant characteristics (MEDIAN (P25–P75), [MIN–MAX]).

|  | Time | Groups |  |  |  | P-value |
| --- | --- | --- | --- | --- | --- | --- |
|  |  | Group 1 | Group 2 | Group 3 | Group 4 |  |
| Sample size (N) | - | 40 | 47 | 76 | 38 | - |
| Sex (F/M) | - | 27/13 | 30/17 | 49/27 | 21/17 | - |
| Age (yrs) | T1 | 76 (71-81) | 80 (68-83) | 77 (73-82) | 78 (71-84) | 0.67 |
| Days since sur. (days) | T1 | 3 (3-4)<br>[2-13] | 28 (23-35)<br>[15-41] | 93 (62-113)<br>[43-178] | 333 (227-355)<br>[189-366] | - |
|  | T2 | 169 (167-182)<br>[148-211] | 205 (185-219)<br>[173-248] | 261 (239-293)<br>[203-376] | 511 (411-526)<br>[348-561] | - |
|  | T3 | 359 (351-370)<br>[340-395] | 380 (368-390)<br>[357-427] | 442 (416-474)<br>[388-559] | 688 (599-707)<br>[538-738] | - |
| Height (cm) | T1 | 168<br>(160-175) | 170<br>(161-180) | 168<br>(162-175) | 168<br>(163-180) | 0.89 |
| Mass (kg) | T1 | 67 (61-75) | 74 (64-83) | 66.8 (56-82) | 71 (59-77) | 0.51 |
|  | T3 | 65 (60-77) | 73 (60-83) | 70 (57-80) | 71 (60-81) | 0.66 |
|  | P-value | 0.76 | 0.76 | 0.29 | 0.15 | - |
| BMI (kg/m<sup>2</sup>) | T1 | 24.0 (21.9-26.7) | 24.8 (22.4-27.5) | 23.7(21.3-27.3) | 23.8(21.8-26.3) | 0.73 |
|  | T3 | 23.8 (21.7-26.6) | 24.2 (22.4-26.8) | 23.7 (21.1-26.9) | 25.2 (21.9-26.9) | 0.81 |
|  | P-value | 0.76 | 0.79 | 0.29 | 0.15 | - |

N: number, F: female, M: male, BMI: body mass index, T1: Baseline, T2: 6-month follow-up, T3: 12-month follow-up. Note: P-values were adjusted using Benjamini & Hochberg's method [31], [32].

The AvAcc and IG of participants during the three assessments are presented in Table II. Median values of AvAcc and IG did not differ significantly between time points except for IG in the extended recovery group (Table II).

**TABLE II.** Average Acceleration (AvAcc) AND Intensity Gradient (IG) Across different recovery groups (MEDIAN (P25–P75))

| Groups |  | Assessments |  |  | P-value | Effect size |
| --- | --- | --- | --- | --- | --- | --- |
|  |  | T1 | T2 | T3 |  |  |
| 1 | AvAcc (mg) | 20 (6–31) | 17 (9–36) | 24 (11–42) | 0.74 | 0.01 |
|  | IG | -3.1 (-4.7--2.0) | -2.9 (-3.3--2.1) | -2.8 (-3.2--2.1) | 0.13 | 0.07 |
| 2 | AvAcc (mg) | 23 (11–38) | 19 (8–32) | 13 (7–25) | 0.76 | 0.01 |
|  | IG | -2.8 (-3.9--1.6) | -3.0 (-3.6--2.3) | -3.1 (-3.7--2.5) | 0.69 | 0.03 |
| 3 | AvAcc (mg) | 13 (7–24) | 20 (9–34) | 16 (9–28) | 0.18 | 0.03 |
|  | IG | -3.2 (-3.8--2.7)* | -3.0 (-3.4--2.5) | -3.0 (-3.7--2.5)* | **0.05** | **0.05** |
| 4 | AvAcc (mg) | 14 (7–27) | 17 (10–37) | 16 (9–34) | 0.29 | 0.04 |
|  | IG | -3.1 (-3.9--2.7) | -3.0 (-3.5--2.5) | -2.9 (-3.4--2.6) | 0.63 | 0.02 |

T1: Baseline, T2: 6-month follow-up, T3: 12-month follow-up. \* T1 versus T3. Effect size magnitude is color-coded based on the applied statistical test: yellow (negligible) [33] Strength of effect size: yellow (negligible). Note: P-values were adjusted using Benjamini & Hochberg's method [31], [32].

Radar plots of the MX metrics (axes: M1-M90) over the three assessments and for each group are depicted in Fig. 1 and Table B in the appendix. Overall, the median values of MX metrics at T2 and T3 were similar and significantly higher than the T1 assessment for groups 1 to 3. The effect size was larger for group 1 compared to groups 2 and 3. For example, median value of M5 in group 1 at T2 and T3 were 81 mg and 86 mg which was significantly higher than T1 (51 mg) (p <0.001, Partial Eta Squared: 0.31 (medium)). Moreover, as the duration of MX metrics increased, the differences between T1 and follow-ups became smaller. However, the median values of MX metrics were almost similar over the three assessments for group 4.

**Fig. 1.**
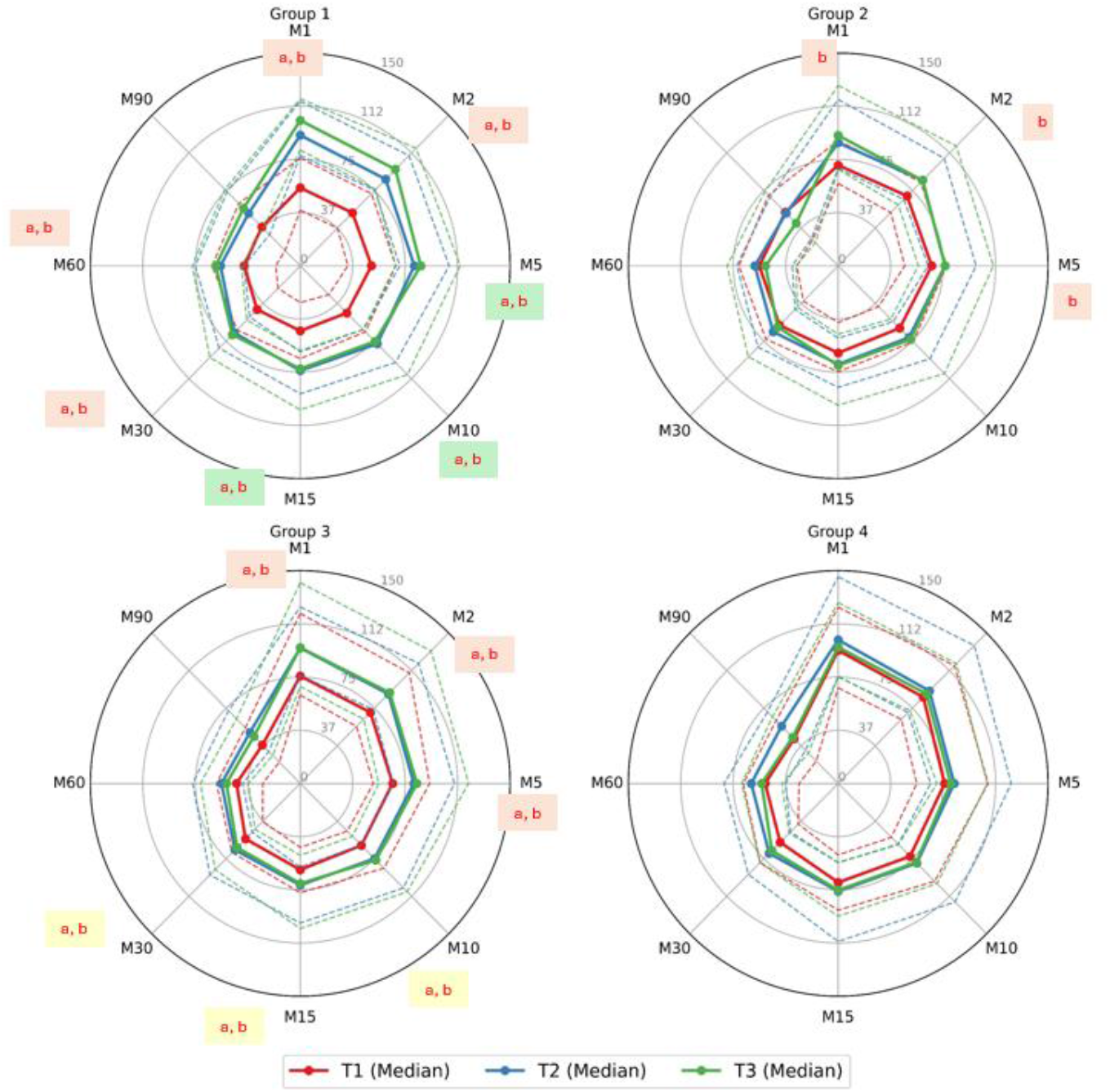
Radar plots illustrating MX metrics (mg). T1: Baseline, T2: 6-month follow-up, T3: 12-month follow-up. Median values: filled line, dashed line: P25 and P75. a) significant differences between T2 and T1, b) significant differences between T3 and T1. Effect size magnitude is colour-coded based on the applied statistical test: medium (green), orange (small), yellow (negligible) [33].

The results of the mobility capacity outcomes (6MinWT, 4MWT and SPPB) at each assessment (T1, T2, and T3) are presented in Table III. Overall, the greatest changes and largest effect sizes were observed in the 6MWT followed by the SPPB and 4MWT across the four groups. More specifically, the distance covered during the 6MinWT increased 41 m in group 1 at T3 compared to T2 (p <0.001, Cohen’s d: 0.92 (large)). The second group walked 72 m and 103 m more at T2 and T3 follow-ups compared to T1 in the 6MinWT, respectively (p<0.001, Partial Eta Squared: 0.63 (large)). The 6MinWT distance increased in group 3 and 4 from T1 to T2 (55 m, 16 m) and T2 to T3 (8 m, 16 m), respectively (p<0.01, Partial Eta Squared: 0.26 (medium), 0,21 (small)). Moreover, the SPPB score increased at both follow-up assessments compared to T1 and the effect size was large among the first two groups (p<0.001, Kendall’s W: 0.77, 0.55 (large)). In group 1, 4MWT speed at T1 was 0.36 m.s^-1^ which increased to 0.86 m.s^-1^ at T2 and 0.91 m.s^-1^ at T3 (p<0.001, Partial Eta Squared: 0.76 (large)). The 4MWT speed also significantly increased in groups 2 and 3 (p<0.001) but did not change in group 4.

**TABLE III.** Mobility capacity tests Across different recovery groups (MEDIAN (P25-P75)).

|  | Groups | Assessments |  |  | P-value | Effect size |
| --- | --- | --- | --- | --- | --- | --- |
|  |  | T1 | T2 | T3 |  |  |
| 6MinWT (m) | Group 1 | - | 379 (282-448) € | 420 (320-487) € | < 0.001 | 0.92 |
|  | N (NA) | 0 (40) | 40 (0) | 37 (3) |  |  |
|  | Group 2 | 255 (167-306) * | 327 (238-453) *€ | 358 (282-477) *€ | < 0.001 | 0.63 |
|  | N (NA) | 45 (2) | 42 (5) | 38 (9) |  |  |
|  | Group 3 | 295 (208-406) * | 349 (272-463) *€ | 357 (254-498) € | < 0.001 | 0.26 |
|  | N (NA) | 74 (2) | 70 (6) | 68 (8) |  |  |
|  | Group 4 | 362 (228-443) * | 378 (258-469) *€ | 394 (285-480) € | 0.009 | 0.21 |
|  | N (NA) | 38 (0) | 37 (1) | 34 (4) |  |  |
| 4MWT (m/s) | Group 1 | 0.36 (0.22-0.42) * | 0.86 (0.73-1.07) * | 0.91 (0.74-1.06) * | < 0.001 | 0.76 |
|  | N (NA) | 40 (0) | 40 (0) | 40 (0) |  |  |
|  | Group 2 | 0.66 (0.52-0.82) * | 0.85 (0.61-0.95) *€ | 0.86 (0.68-1.03) *€ | < 0.001 | 0.37 |
|  | N (NA) | 47 (0) | 47 (0) | 45 (2) |  |  |
|  | Group 3 | 0.75 (0.58-0.95) * | 0.86 (0.67-1.12) * | 0.89 (0.68-1.11) * | < 0.001 | 0.14 |
|  | N (NA) | 76 (0) | 74 (2) | 74 (2) |  |  |
|  | Group 4 | 0.90 (0.64-1.16) | 0.92 (0.69-1.17) | 0.99 (0.74-1.16) | 0.427 | 0.04 |
|  | N (NA) | 38 (0) | 37 (1) | 37 (1) |  |  |
| SPPB (/12) | Group 1 | 4 (2-5) * | 10 (8-11) * | 10 (8-11) * | < 0.001 | 0.77 |
|  | N (NA) | 40 (0) | 40 (0) | 40 (0) |  |  |
|  | Group 2 | 5 (4-8) * | 8 (7-10) * | 9 (6-11) * | < 0.001 | 0.55 |
|  | N (NA) | 47 (0) | 47 (0) | 45 (2) |  |  |
|  | Group 3 | 8 (6-10) * | 9 (7-11) * | 9 (7-11) * | < 0.001 | 0.20 |
|  | N (NA) | 76 (0) | 74 (2) | 74 (2) |  |  |
|  | Group 4 | 9 (6-11) | 10 (6-11) | 10 (6-12) | 0.044 | 0.11 |
|  | N (NA) | 38 (0) | 37 (1) | 37 (1) |  |  |

6MinWT: 6-min walking test distance, 4MWT: 4-meter walking test, SPPB: Short Physical Performance Battery, N: sample size, NA: Not Available, T1: Baseline, T2: 6-month follow-up, T3: 12-month follow-up. Note: Significant differences are marked in bold. \* T1 versus T2, T3, € T2 versus T3.
Effect size magnitude is color-coded based on the applied statistical test: large (purple), medium (green), orange (small), yellow (negligible) [33]. P-values were adjusted using Benjamini & Hochberg's method [31], [32].

Mobility perception changes, derived from the LLFDI questionnaire, are depicted in radar plots (Fig. 2, Table C in appendix). The axes displayed in these plots represent the seven LLFDI’s domains. Overall, LLFDI’s changes were significantly different only for groups 2 and 3. In group 2, Basic and Advanced Lower Extremity median values were 51.3 and 11.4 at T1, respectively. They significantly increased at T2 (63.8, 42.8) and T3 (68.6, 49.7) (p <0.001, Kendall’s W: 0.51, 0.4 (large, medium)). The median value of Social Role increased 8.1 and 6.8 points at T2 and T3 in group 2 (p<0001, Kendall’s W: 0.31 (medium)). In group 2, changes in Instrumental Role, and Management Role, and Personal Role (PR) scores were also significant (p<0.01), although the effect sizes were small. The increase in LLFDI scores followed a similar pattern in group 3 with lower effect size.

**Fig. 2.**
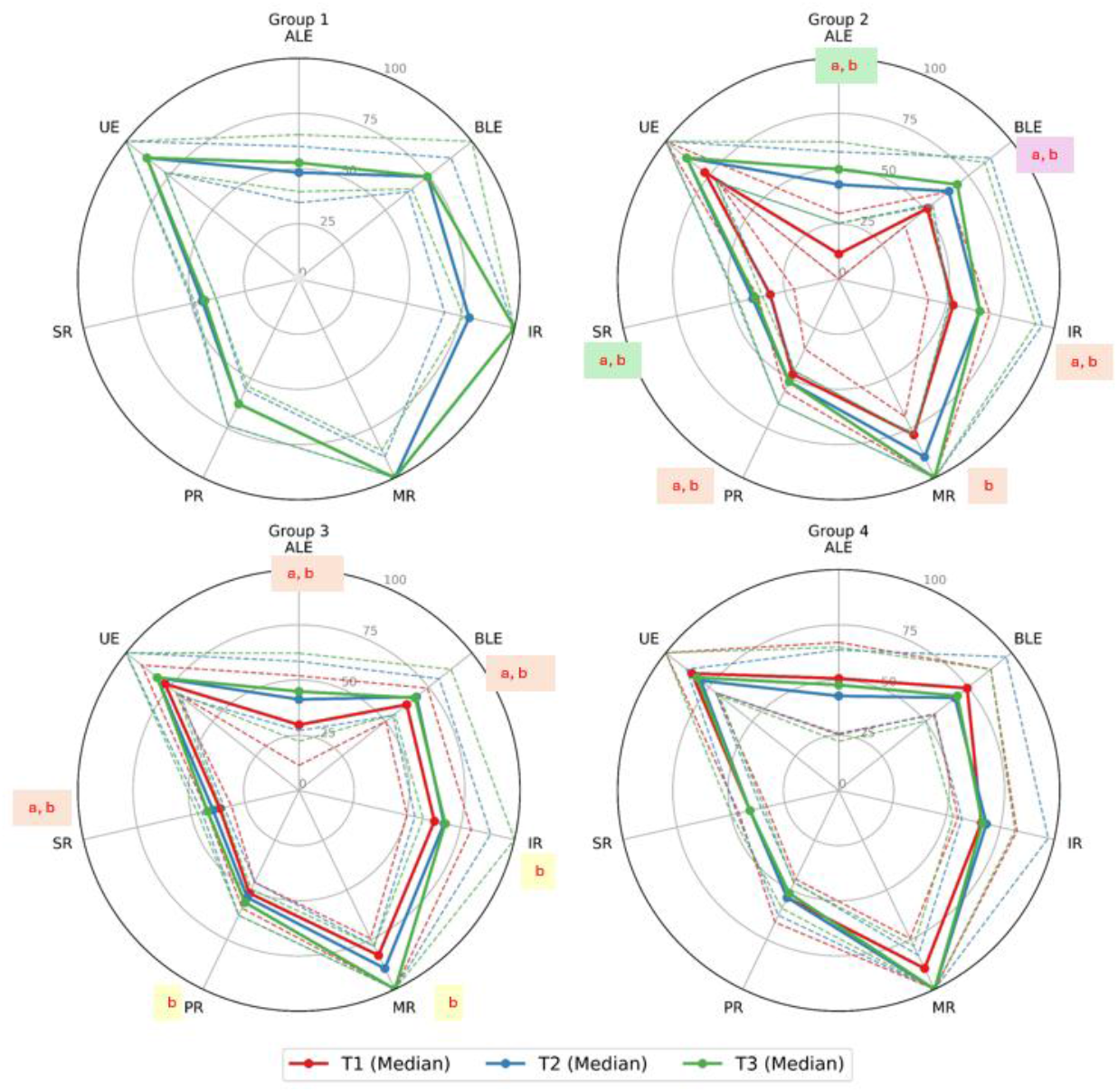
Radar plots illustrating LLFDI’s domains: Advanced Lower Extremity (ALE), Basic Lower Extremity (BLE), Instrumental Role (IR), and Management Role (MR), Personal Role (PR), Social Role (SR), Upper Extremity (UE). T1: Baseline, T2: 6-month follow-up, T3: 12-month follow-up. Median values: filled line, dashed line: P25 and P75. a) significant differences between T2 and T1, b) significant differences between T3 and T1. Effect size magnitude is color-coded based on the applied statistical test: large (purple), medium (green), orange (small), yellow (negligible) [33].

The heat map plots presented in Fig. 3 show the correlations between clinical tests (y-axis) and cut-point free metrics (x-axis) for each assessment and group separately. Overall, we found a high correlation between mobility capacity tests and short-duration MX metrics (M1-M15). Particularly, 6MinWT showed the strongest correlation, followed by SPPB and 4MWT. Strong correlations were observed between short-duration MX metrics and LLFDI’s Function component (Advanced Lower Extremity, Basic Lower Extremity). Within the LLFDI’s Disability component, the Instrumental Role and Social Role domains showed the highest correlation with short-duration MX metrics.

**Fig. 3.**
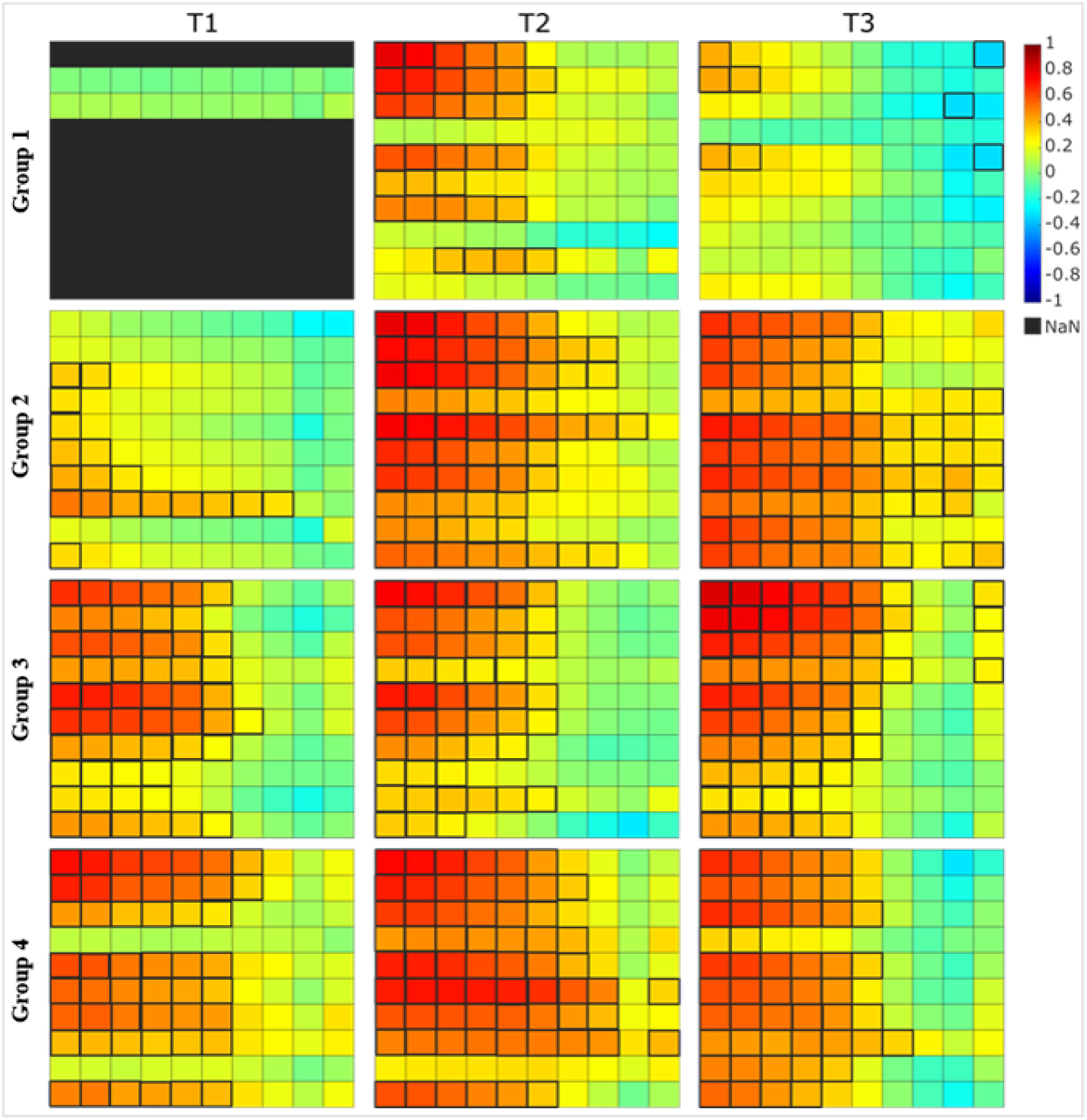
Heat map plots illustrating the level of association (r_s_ value) between cut-point free metrics and all clinical tests. For all the plots: Y axis from top to bottom: 1) 6MinWT, 2) 4MWT, 3) SPPB, LLFDI’s domains including: 4) Upper Extremity, 5) Advanced Lower Extremity, 6) Basic Lower Extremity, 7) Instrumental Role, 8) Management Role, 9) Personal Role, 10) Social Role. X axis from left to right: cut-point. free metrics: 1) M1, 2) M2, 3) M5, 4) M10, 5) M15, 6) M30, 7) M60, 8) M90, 9) AcAcc, 10) IG. **Black border** means significant correlations.

Group 1 did not show a constant association pattern over the three assessments. However, their heat map plots at T2 showed more similarity with the participants in the other groups with large correlation between mobility capacity tests, LLFDI’s Advanced Lower Extremity domain, and M1-M15 (i.e., r_s_ between 6MinWT and M1: 0.79 (95% CI: 0.64-0.89)). In group 2 at T1, the relationship between cut-point free metrics and clinical assessments were generally weak. The strongest correlation was 0.51 (95% CI: 0.26-0.70) corresponding to the LLFDI’s Management Role (Limitation Dimension) and M1 (p<0.001). However, the strength of correlations increased within group 2 at T2 and T3. For example, at T2 the correlations (95% CI) between 4MWT and M1 and between 4MWT and M15 were 0.73 (0.55-0.84) and 0.55 (0.31-0.72), respectively.

Heat map plots in groups 3 and 4 showed more stable association patterns over the three assessments. The highest correlation was 0.81 (95% CI: 0.72-0.88) between the 6MinWT and M1 in group 3 at T3 (p<0.001). The correlations between SPPB and M1 and between SPPB and M5 were 0.68 (0.53-0.78) and 0.62 (0.45-0.74) at T3 in group 4, respectively (p<0.001). Correlations between the Instrumental Role domain of LLFDI (Limitation Dimension) and MX metrics (e.g., M1) were higher than the Management Role domain (e.g., in group 4 at T3: 0.56 (95% CI: 0.3-0.75) versus 0.46 (95% CI: 0.16-0.68)). In LLFDI’s Frequency Dimension, Social Role domain had higher correlations with MX metrics compared to Personal Role among most participants and across assessments. For instance, in group 4, the correlations of Social Role and Personal Role with M5 were 0.59 (95% CI: 0.34-0.77, p<0.01) and 0.30 (95% CI: -0.02-0.57, p=0.07) at T2, respectively.

## IV. Discussion

### A. Change in daily PA, mobility capacity, and mobility perception

AvAcc (PA volume) and IG (PA intensity) showed an increasing trend at T3 compared to T1, even though not statistically different. The minimal clinically important difference (MCID) represents the smallest detectable meaningful change for patients, clinicians, and researchers. Rowlands and colleagues used three approaches to estimate MCID in mg for inactive adults based on wrist-worn accelerometers measurements [24]. First, an increase of 1 mg for AvAcc was equivalent to 500 steps/day, which is associated with a 8-9% decreased risk of all-cause mortality in older women [24], [36]. Second, a 0.8-1 mg AvAcc increase was equivalent to substituting 5-6 min spent inactive (e.g., sitting) with 5-6 min of brisk walking [21], [24]. Third, they directly assessed an increase of 1 mg AvAcc and health from UK Biobank data. They found that higher AvAcc (per 1 mg) was associated with lower all-cause mortality. Therefore, the 2-4 mg increase in AvAcc between T3 and T1 in our participants, to the exception of group 2, can be interpreted as an additional 100-2000 steps/day, or 10-24 min brisk walk throughout the day, which might reduce all-cause mortality in PFF patients. However, acceleration signals depend on the body sensor’s location. Migueles and colleagues compared AvAcc between dominant wrist, non-dominant wrist, and right hip placements among young adults. [37] Their findings showed that the AvAcc of hip-worn sensor was almost half the dominant wrist worn sensor. Hence, 2-4 mg increase of AvAcc recorded by a lower-back sensor, as is the case in our study, might translate to more than 100-2000 daily steps with higher health benefits.

The increase trend in IG indicates that they spent more time across mid and high range of PA intensity [21], [25]. It might indicate a more diverse movement capability, likely due to the recovery process. Moreover, Schwendinger and colleagues found that higher daily PA intensity distribution (IG) was more closely related to reduced mortality risk than PA volume (AvAcc) [38].

Additionally, MX metrics (proxy for time-related intensity of daily PA), particularly short-duration MX (i.e., M1-M15), increased significantly at T2 and T3 compared to T1 among all groups except group 4. This finding aligned with previous studies that demonstrated the suitability of short-term MX metrics for detecting group differences [19], [27]. Group 4 was at a late stage of recovery since T1, which might explain the stability of the MX metrics across assessments. However, the recovery stage in group 1 shifted from acute at T1 to long-term recovery at T3. While group 2 and 3 at T1 went from post-acute and extended recovery to long-term recovery at T3, respectively. The large effect size of short-duration MX metrics in group 1 indicates their high sensitivity to change, which might be due to a broader spectrum of recovery stage across assessments.

Among mobility capacity tests, the 6MinWT demonstrated the highest sensitivity to change, followed by the SPPB and 4MWT. This finding agreed with the study by Gill suggesting that more challenging tests, such as long-distance corridor walk, decrease the potential ceiling effects in longitudinal studies [39]. Furthermore, mobility capacity tests were more sensitive to change over three assessments in groups 1 and 2 compared to 3 and 4. This could reflect broader recovery trajectory in the first two groups, as they transitioned from high disabilities at T1 to lower disability at T2 and T3 due to the longer duration of post-surgery. Perera and colleagues concluded that MCID of these three tests are 20 meters for 6MinWT, 0.05 m.s^-1^ for 4MWT, and 0.5 points for SPPB; and that substantial meaningful changes were 50 meters for 6MinWT, 0.10 m.s^-1^ for 4MWT, and 1 point for SPPB among older adults [40], [41], [42]. Our findings first showed that nearly all the participants reached at least one of these MCID thresholds. Second, the improvement in mobility capacity across all three tests reached substantial thresholds between T1 to T2 and T3 particularly in the first two groups. Third, the distance covered during the 6MinWT increased significantly between T2 and T3, exceeding 20 m in groups 1 and 2. Lastly, group 4 at T1 and all participants at T2 and T3 reached >300 m in the 6MinWT indicating sufficient ability for community ambulation [43], [44].

LLFDI results showed significant mobility perception change for group 2 and 3 between T1 and follow-up assessments (T2 and T3). These improvements were observed in the Function component for both the Basic Lower Extremity domain (fundamental movements like standing and stooping) and the Advanced Lower Extremity domain (endurance and high-ability activities). The corresponding MCID threshold for substantial changes were 6 and 9 points for these domains, respectively [45]. Group 2 met both thresholds with greater sensitivity to change than group 3, which only reached MCID for the Advanced Lower Extremity domain. Additionally, the Disability component scores improved significantly. Notably, the Social Role domain demonstrated the greatest sensitivity to change. Based on our knowledge, there is no available MCDI for the LLFDI Disability component.

### B. Association between daily PA, mobility capacity, and mobility perception

Overall, heat map plots showed moderate to strong positive correlation between mobility capacity tests (6MinWT, SPPB, 4MWT) and short-duration MX metrics (M1-M15) over three assessments. It could suggest that short duration and high intensity daily PA are of a similar nature to mobility capacity tests. This finding aligned with a previous study [27] which found a similar relationship between lower limb mobility capacity tests and MX metrics among individuals recovering from PFF.

The correlations between LLFDI’s Basic and Advanced Lower Extremity LLFDI scores and short-duration MX metrics were positive and strong. It could be interpreted as the perceived lower limb functional mobility (e.g., standing, stepping, walking, running) of participants and their high intensity daily PA having a direct relationship. Similarly, the high positive correlation between the Instrumental and Social Role domains of LLFDI scores with short-duration MX metrics could suggest that individuals with higher daily PA intensity had less perceived limitation for performing daily PA and higher perceived frequency for social activities.

Weak correlations between the clinical tests and cut-point free metrics in groups 1 and 2 at T1 might be related to their early recovery stage (acute and post-acute). The higher and consistent correlations observed in group 3 and 4 at T1 and across all participants at T2 and T3 might indicate greater recovery stability and more robust relationship between clinical assessments and daily PA (short-duration MX metrics). These findings were aligned with Newman and colleague who found that the outcomes of long-distance corridor walking were strongly associated with mobility disabilities [46]. The heat map plot for group 1 at T3 was exceptional and did not follow the same patterns as the other groups. The lower correlations might be related to change in rehabilitation program or seasonal conditions from T2 to T3 and need further investigation.

### C. Limitations

The limitations of this study were a lack of information about contextual factors (e.g., environmental, motivation, activity type); mobility perception assessment by LLFDI interview and their vulnerability to external influences such as examiner; and recording movement artefacts by body worn accelerometers. Despite these limitations, this study provides meaningful insights into the high sensitivity to change of short-duration MX metrics to capture daily PA change, as well as for the 6MinWT and three LLFDI’s domains (Basic Lower Extremity, Advanced Lower Extremity, Social Role). This was particularly salient for groups 1 and 2, which moved from very early recovery stage to late recovery stage over the assessments. Further longitudinal studies with comprehensive contextual factors (e.g., daily activity type, rehabilitation program between each data assessment) are essential to better understand the relationships between daily PA intensities and clinical tests.

## V. Conclusion

Among cut-point free metrics, short-duration MX metrics (i.e., M1-M15) captured changes in daily PA intensity with higher sensitivity to change among older adults recovering from PFF than long-duration MX metrics. The 6MinWT demonstrated the highest sensitivity, and participants achieved substantial improvement across all three assessments. To reduce clinical burden and maximize efficiency, clinicians can only focus on the three LLFDI domains that showed the largest significant changes (Basic Lower Extremity, Advanced Lower Extremity, Social Role). Sensitivity of these changes was higher in individuals who had a wider recovery spectrum (groups 1 and 2). The consistent, positive, moderate to strong correlations between the mobility capacity tests and short-duration MX metrics observed in late recovery stage at T1 and all participants at T2 and T3 (excluding group 1 at T3) suggest a similar nature of high intensity and short-duration activities. Therefore, MX metrics have potential for clinical applications and remote monitoring. Moderate to strong positive correlations between four LLFDI domains (Basic Lower Extremity, Advanced Lower Extremity, Personal Role, Social Role) and short-duration MX metrics indicate a direct link between higher perceived lower limb function, lower levels of perceived disability in daily and social activities, and higher daily PA intensities.

## Data Availability

All data produced in the present study are available upon reasonable request to the correspond author, subject to formal review and approval by the Data Access Committee.

## Acknowledgments

The authors thank Quentin Le Cornu biostatistician at UCD CSTAR for his valuable statistical assistance and reviewing the manuscript. The authors also would like to thank UCD physiotherapist students Hugh O’Brien, Hugh Hogan, Calum Dowling, and Deirdre B. Earls for their contribution to data interpretation. The authors also gratefully acknowledge all participants for their time and contributions. Full membership of the Mobilise-D consortium is available on the website (URL: https://mobilise-d.eu/about-us/, accessed on 28 September 2023).

